# Prognostic Value of Mean Platelet Volume in Septic Shock: A Retrospective Cohort Study

**DOI:** 10.64898/2026.05.29.26354453

**Authors:** Fernando Trujillo-Vega, Phabel A. Lopez-Delgado

## Abstract

**Background:** Mean platelet volume (MPV) is a simple, low-cost biomarker that reflects platelet activation. Its prognostic value in septic shock remains controversial. We aimed to determine whether MPV at intensive care unit (ICU) admission is associated with hospital mortality in patients with septic shock.

**Methods:** Retrospective cohort study of consecutive adults with septic shock (Sepsis-3 criteria) admitted to a single ICU. MPV, severity scores (SOFA, APACHE II, SAPS II), procalcitonin, and clinical data were collected. The primary outcome was in-hospital mortality. Spearman correlation, univariate and multivariate logistic regression (with Firth’s correction), ROC curves, and subgroup analyses were performed.

**Results:** Fifty-eight patients were included; mortality was 58.6%. MPV did not differ between non-survivors and survivors (13.09 *±* 1.37 vs. 12.66 *±*1.45 fL, *p* = 0.259). MPV showed a weak correlation with procalcitonin (*ρ* = 0.394, *p* = 0.002) but not with severity scores. In multivariate analysis adjusting for age, sex, SOFA and comorbidity count, MPV was not an independent predictor of mortality (OR 1.075, 95% CI 0.682–1.755, *p* = 0.749). The area under the ROC curve for MPV was 0.598 (95% CI 0.444–0.752), significantly lower than that of SOFA (0.837) and procalcitonin (0.836). Sub-group analyses showed no significant association between MPV and mortality in any stratum.

**Conclusions:** In this cohort of septic shock patients, MPV at ICU admission was not associated with hospital mortality and had poor discriminative ability. Widely used severity scores and procalcitonin remain superior prognostic markers. MPV should not be used as a prognostic tool in septic shock.

**Highlights:**

1. Retrospective cohort study of 58 patients with septic shock (Sepsis-3 criteria).
2. MPV at ICU admission was not associated with hospital mortality (OR 1.075, 95% CI 0.682–1.755, *p* = 0.749).
3. MPV had poor discriminative ability (AUC 0.598), significantly inferior to SOFA (0.837) and procalcitonin (0.836).
4. Subgroup analyses (age, sex, SOFA score) found no significant effect of MPV on mortality.
5. MPV should not be used as a prognostic biomarker in septic shock.

## 2. Introduction

Sepsis is defined as life-threatening organ dysfunction caused by a dys-regulated host response to infection [1]. Its most severe form, septic shock, is characterised by persistent hypotension requiring vasopressors and a serum lactate level > 2mmol/L despite adequate fluid resuscitation [1]. Despite advances in critical care, mortality rates remain high, ranging from 11% to 55% depending on the population studied [2, 3].

Early identification of patients at high risk of death is crucial to guide therapy and allocate resources. Several scoring systems (SOFA, APACHE II, SAPS II) have been developed and validated [4], but they require multiple clinical and laboratory variables. Simple, inexpensive, and readily available biomarkers are needed, especially in resource-limited settings [3, 5].

Platelets play a central role in sepsis pathophysiology, being activated and consumed during the inflammatory response [6, 7]. Mean platelet volume (MPV) is a routine haematological parameter that reflects platelet size. Larger platelets are more reactive, contain more pro-thrombotic granules, and are associated with adverse cardiovascular events [8]. In sepsis, elevated MPV has been proposed as a marker of platelet activation and a potential predictor of mortality [9, 10].

However, evidence is conflicting. Some studies have found that higher MPV is associated with increased mortality in septic patients [9, 11], whereas others reported no independent association after adjustment for disease severity [10]. Moreover, most investigations have included mixed populations (sepsis and septic shock) and have not consistently adjusted for key confounders such as severity scores. Consequently, the specific prognostic value of MPV in pure septic shock cohorts remains unclear.

Therefore, we conducted this retrospective cohort study to determine whether MPV measured at ICU admission is independently associated with in-hospital mortality in adult patients with septic shock.

## 3. Methods

The Graphical Abstract with the entire process is shown in Figure 1.

**Figure 1:**
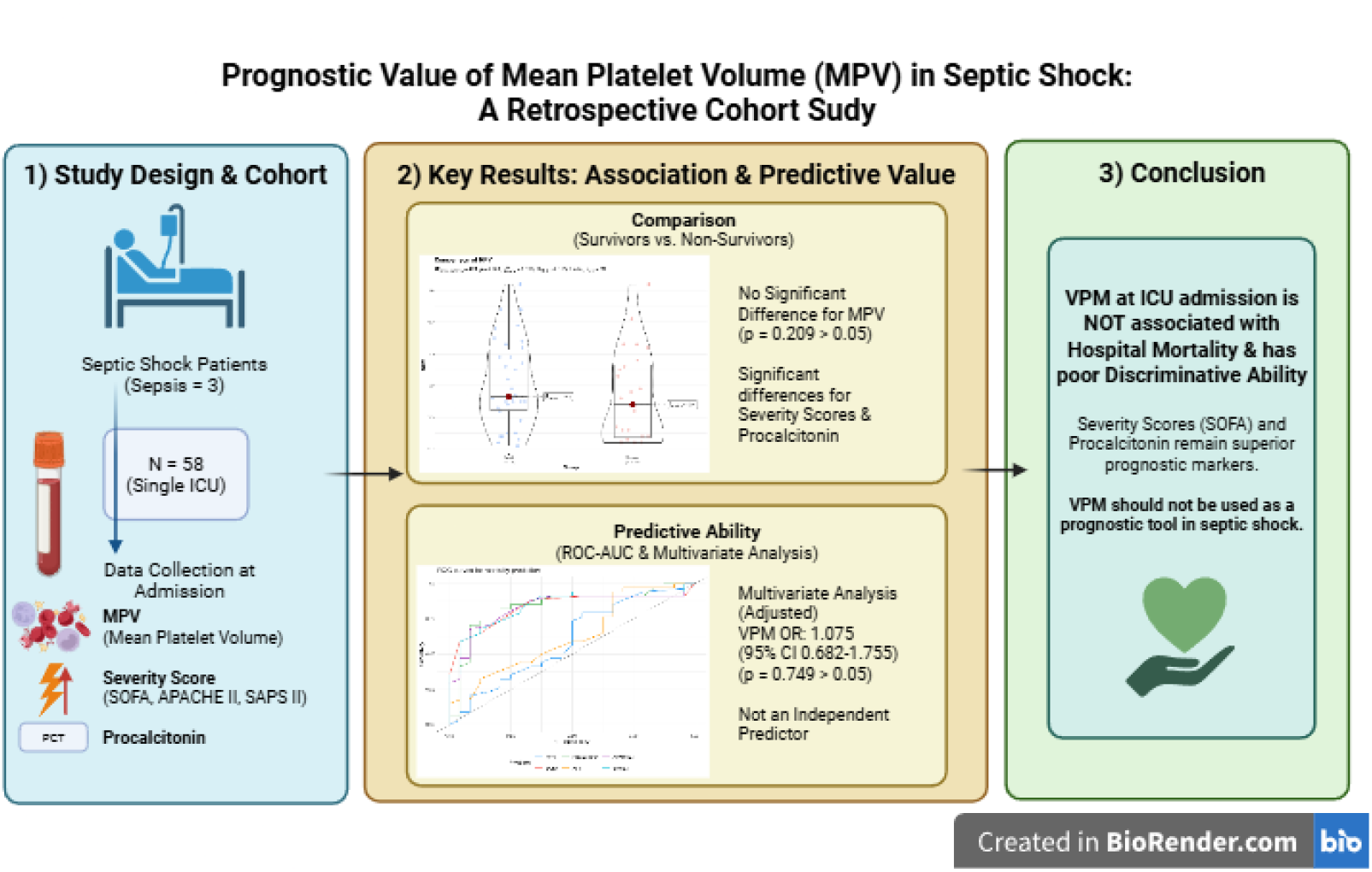
Graphical abstract. Patients with septic shock (n=58) were admitted to the ICU. Mean platelet volume (MPV) measured at admission did not differ significantly between survivors and non-survivors (p=0.259). MPV had poor discriminative ability (AUC=0.598), whereas SOFA (AUC=0.837) and procalcitonin (AUC=0.836) showed good performance. The figure summarizes the main finding: MPV is not a prognostic marker in septic shock. Made with *BioRender*.

### 3.1. Study design and setting

This was a retrospective cohort study conducted at the adult intensive care unit (ICU) of the Hospital Regional “General Ignacio Zaragoza”, ISSSTE, Mexico City. The study was approved by the institutional ethics committee (RPI-HRGIZ-046-2025). Written informed consent was obtained from all participants.

### 3.2. Participants

All consecutive adults (≥ 18 years) admitted to the ICU between [dates] with a diagnosis of septic shock according to the Sepsis-3 criteria [1] were eligible for inclusion. Exclusion criteria were: pregnancy, known immuno-suppressive therapy, HIV infection, active haematological malignancy, death within the first 24 hours before MPV measurement, or missing MPV data.

### 3.3. Data collection

From electronic medical records we extracted: age, sex, comorbidities, admission diagnosis (infection focus), first available MPV within 24 hours of ICU admission, procalcitonin (PCT), and the following severity scores: Sequential Organ Failure Assessment (SOFA), Acute Physiology and Chronic Health Evaluation II (APACHE II), and Simplified Acute Physiology Score II (SAPS II). The primary outcome was all-cause in-hospital mortality.

### 3.4. Statistical analysis

Continuous variables were expressed as mean *±* standard deviation or median (interquartile range) as appropriate, and categorical variables as frequencies (percentages). Comparisons between survivors and non-survivors were performed using the Mann–Whitney *U* test or chi-square test.

Spearman correlation was used to evaluate the relationship between MPV and selected continuous variables (SOFA, APACHE II, SAPS II, procalcitonin, age).

Logistic regression was employed to assess the association between MPV and mortality. Univariate models were fitted for each predictor. Because complete separation was encountered (perfect prediction by some variables), we used Firth’s bias-reduced logistic regression (‘logistf’ package) [10]. The multivariate model included MPV, age, sex, SOFA, and comorbidity count (number of pre-existing diseases) based on clinical relevance and availability.

Receiver operating characteristic (ROC) curves were constructed for MPV, SOFA, procalcitonin, APACHE II, SAPS II, and age. The area under the curve (AUC) was computed with 95% confidence intervals (CI). The DeLong test was used to compare the AUCs of MPV and SOFA.

Subgroup analyses were performed to examine whether the effect of MPV on mortality differed by age (≥ 65 vs *<* 65 years), sex, and SOFA (≥ 10 vs *<* 10). Interaction terms were not formally tested due to limited sample size.

All analyses were performed in R software (version 4.5.3), with packages ‘dplyr’, ‘ggplot2’, ‘pROC’, and ‘logistf’. And the ggstatsplot package for exploratory between-group comparisons [12]. A two-sided *p <* 0.05 was considered statistically significant.

### 3.5. Ethics approval

The study was approved by the institutional research and ethics committee of the Hospital Regional “General Ignacio Zaragoza”, ISSSTE (registration number **RPI-HRGIZ-046-2025**). Because of its retrospective, observational design, the study was not registered in a clinical trial registry. The requirement for informed consent was waived by the committee. The study adheres to the Strengthening the Reporting of Observational Studies in Epidemiology (STROBE) guidelines.

## 4. Results

### 4.1. Patient characteristics

A total of 58 patients with septic shock were included. Overall hospital mortality was 58.6% (34 of 58). Baseline characteristics stratified by survival are shown in Table 1. Non-survivors were significantly older (mean age 66.9± 12.3 vs. 58.6 ± 16.5 years, p = 0.033) and had higher severity scores: SOFA (11.5 ± 2.1 vs. 9.0 ± 1.3, p < 0.001), APACHE II (31.0 ± 7.0 vs. 23.8 ± 3.7, p < 0.001) and SAPS II (66.4±14.0 vs. 50.0±7.4, p < 0.001). Procalcitonin levels were also markedly higher in non-survivors (55.2 ± 34.7 vs. 15.1± 24.2 ng/mL, p < 0.001). Mean platelet volume (MPV) did not differ significantly between survivors and non-survivors (13.09 ± 1.37 vs. 12.66 ± 1.45 fL, p = 0.259). This i shown in Figure 2.

**Table 1:** Baseline characteristics of survivors and non-survivors.

| Variable | Survivors<br>($n = 24$) | Non-survivors<br>($n = 34$) | $p$-value |
| --- | --- | --- | --- |
| Age (years) | $58.6 \pm 16.5$ | $66.9 \pm 12.3$ | 0.033 |
| Male sex, $n$ (%) | 14 (58.3) | 18 (52.9) | 0.890 |
| SOFA score | $9.0 \pm 1.3$ | $11.5 \pm 2.1$ | $< 0.001$ |
| APACHE II score | $23.8 \pm 3.7$ | $31.0 \pm 7.0$ | $< 0.001$ |
| SAPS II score | $50.0 \pm 7.4$ | $66.4 \pm 14.0$ | $< 0.001$ |
| Procalcitonin (ng/mL) | $15.1 \pm 24.2$ | $55.2 \pm 34.7$ | $< 0.001$ |
| MPV (fL) | $12.66 \pm 1.45$ | $13.09 \pm 1.37$ | 0.259 |
| Comorbidity count | $2.17 \pm 0.87$ | $1.76 \pm 0.70$ | 0.056 |

Data are mean ± standard deviation unless otherwise stated.
$p$ -values from Mann–Whitney $U$ test (continuous) or chi-square test (categorical).

**Figure 2:**
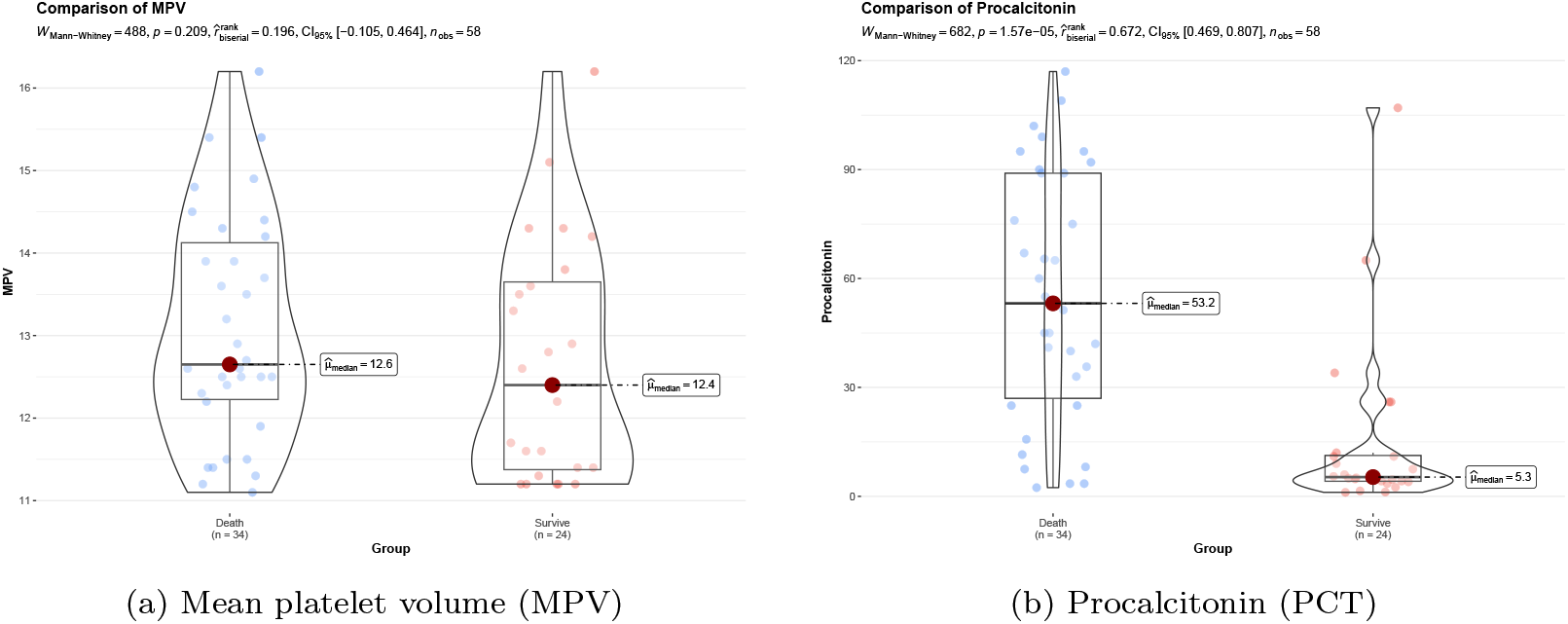
Boxplots of MPV and procalcitonin in survivors and non-survivors. MPV did not differ significantly (*p* = 0.259), whereas procalcitonin was markedly higher in non-survivors (*p <* 0.001).

### 4.1.1. Correlation of MPV with clinical variables

Spearman correlation coefficients between MPV and selected variables are presented in Table 2. MPV showed a weak, non-significant correlation with SOFA (*ρ* = 0.243, *p* = 0.066), APACHE II (*ρ* = 0.242, *p* = 0.067) and SAPS II (*ρ* = 0.243, *p* = 0.066). A moderate positive correlation was observed with procalcitonin (*ρ* = 0.394, *p* = 0.002). No correlation was found with age (*ρ* = − 0.065, *p* = 0.630). The scatter plot of MPV versus procalcitonin is shown in Figure 3.

**Table 2:** Spearman correlations of MPV with selected variables.

| Variable | $\rho$ | 95% CI | $p$-value |
| --- | --- | --- | --- |
| SOFA | 0.243 | 0.084 – 0.395 | 0.066 |
| APACHE II | 0.242 | 0.083 – 0.394 | 0.067 |
| SAPS II | 0.243 | 0.084 – 0.395 | 0.066 |
| Procalcitonin | 0.394 | 0.220 – 0.552 | 0.002 |
| Age | −0.065 | −0.315 – 0.190 | 0.630 |

**Figure 3:**
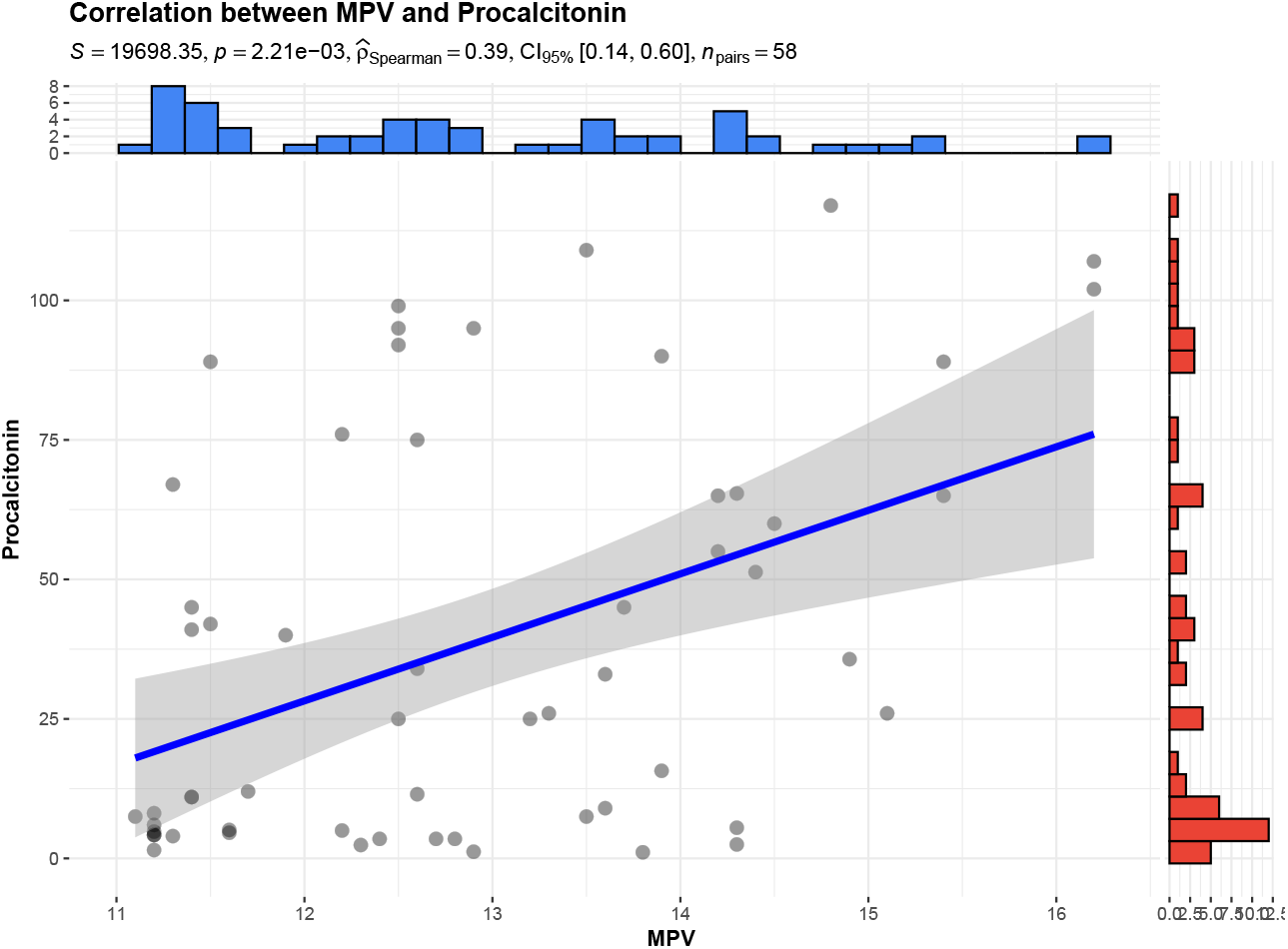
Scatter plot of MPV versus procalcitonin with Spearman correlation coefficient (*ρ* = 0.394, *p* = 0.002). The solid line represents the regression line.

### 4.1.2. Predictors of mortality

Univariate logistic regression (Table 3) identified age, SOFA, APACHE II, SAPS II and procalcitonin as significant predictors of mortality (all *p <* 0.05). MPV was not significantly associated with mortality (OR 1.235, 95% CI 0.854–1.838, *p* = 0.273).

**Table 3:** Univariate logistic regression for mortality.

| Predictor | OR | 95% CI | $p$-value |
| --- | --- | --- | --- |
| MPV (per fL) | 1.235 | 0.854 – 1.838 | 0.273 |
| Age (per year) | 1.040 | 1.002 – 1.084 | 0.046 |
| SOFA (per point) | 2.015 | 1.441 – 3.099 | $< 0.001$ |
| APACHE II (per point) | 1.269 | 1.119 – 1.515 | 0.001 |
| SAPS II (per point) | 1.139 | 1.070 – 1.237 | $< 0.001$ |
| Procalcitonin (per ng/mL) | 1.044 | 1.021 – 1.074 | $< 0.001$ |
| Comorbidity count | 0.526 | 0.257 – 1.023 | 0.065 |

In the multivariate model (Table 4) adjusting for age, sex, SOFA and comorbidity count, only SOFA remained independently associated with mortality (OR 1.873 per point, 95% CI 1.333–2.905, *p <* 0.001). MPV was not significant (OR 1.075, 95% CI 0.682–1.755, *p* = 0.749).

**Table 4:** Multivariate logistic regression model for mortality.

| **Predictor** | **OR** | **95% CI** | **p-value** |
| --- | --- | --- | --- |
| MPV (per fL) | 1.075 | 0.682 – 1.755 | 0.749 |
| Age (per year) | 1.042 | 0.997 – 1.100 | 0.075 |
| Sex (male vs. female) | 0.976 | 0.260 – 3.743 | 0.970 |
| SOFA (per point) | 1.873 | 1.333 – 2.905 | < 0.001 |
| Comorbidity count | 0.581 | 0.249 – 1.288 | 0.175 |

Model adjusted for all variables shown.

### 4.1.3. ROC analysis

Area under the ROC curves (AUC) for mortality prediction are shown in Table 5. MPV had poor discriminative ability (AUC 0.598, 95% CI 0.444–0.752). In contrast, SOFA (AUC 0.837, 95% CI 0.731–0.943), procalcitonin (AUC 0.836, 95% CI 0.723–0.949), APACHE II and SAPS II all showed good to excellent performance. The DeLong test confirmed that SOFA significantly outperformed MPV (*p* = 0.005). The ROC curves are displayed in Figure 4. The DeLong test confirmed that SOFA significantly outperformed MPV (*p* = 0.005).

**Table 5:** Area under the ROC curve (AUC) for mortality prediction.

| **Predictor** | **AUC** | **95% CI** | **p (vs. MPV)<sup>a</sup>** |
| --- | --- | --- | --- |
| MPV | 0.598 | 0.444 – 0.752 | – |
| SOFA | 0.837 | 0.731 – 0.943 | 0.005 |
| Procalcitonin | 0.836 | 0.723 – 0.949 | 0.007 |
| APACHE II | 0.834 | 0.724 – 0.944 | 0.008 |
| SAPS II | 0.848 | 0.746 – 0.950 | 0.004 |
| Age | 0.626 | 0.478 – 0.774 | 0.782 |

<sup>a</sup> DeLong test comparing each predictor with MPV.

**Figure 4:**
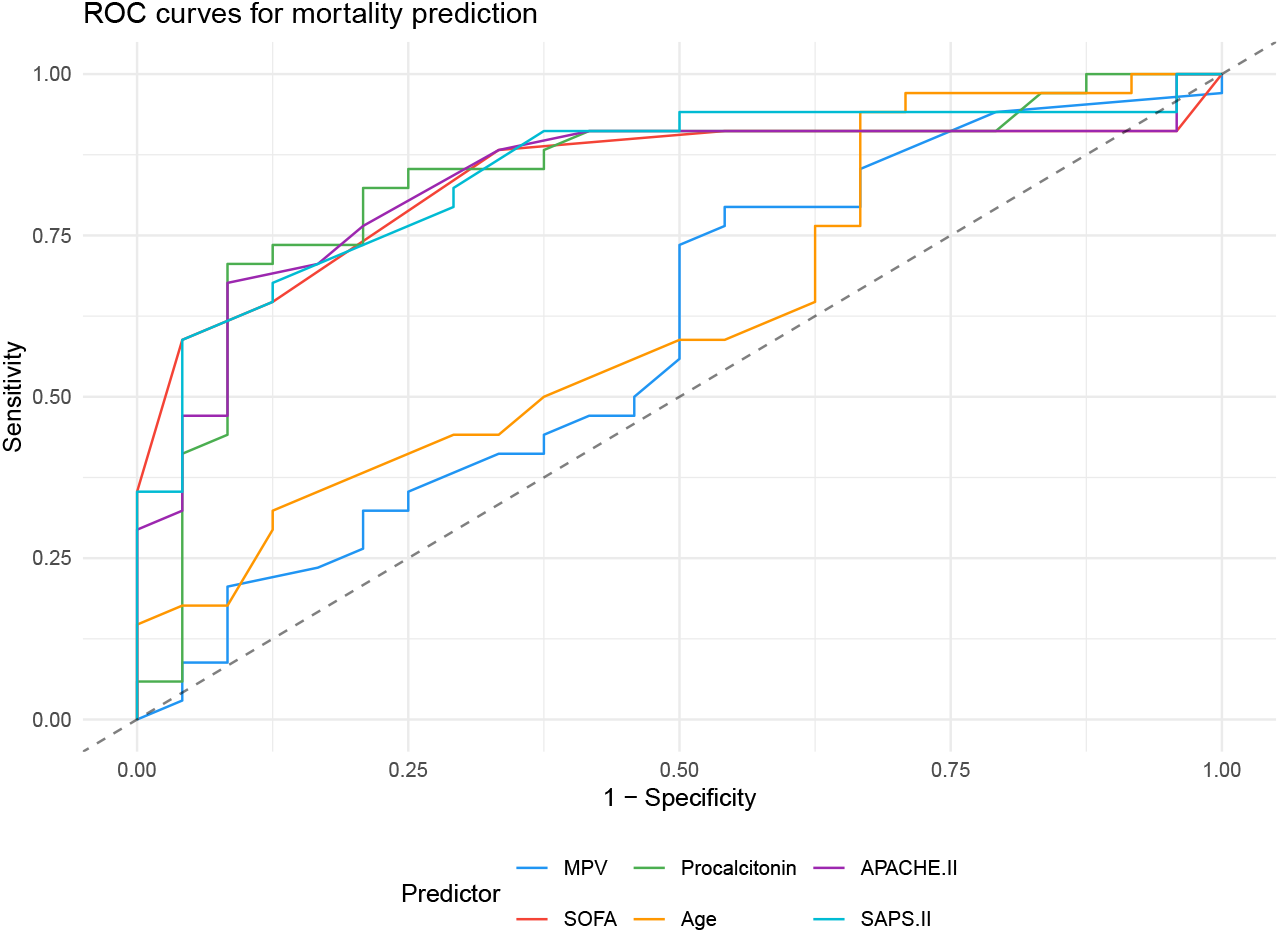
ROC curves for mortality prediction. AUC values are reported in Table 5. MPV (dashed line) shows poor discriminative ability (AUC 0.598) compared to SOFA (0.837), procalcitonin (0.836), APACHE II (0.834), and SAPS II (0.848).

### 4.1.4. Subgroup analysis

We examined the effect of MPV on mortality across patient subgroups (Table 6). No subgroup showed a significant association (all *p* > 0.05). A trend towards higher mortality with increasing MPV was observed in patients younger than 65 years (OR 1.83, 95% CI 1.01–3.82, *p* = 0.066), but this did not reach statistical significance. A forest plot of the subgroup analysis is presented in Figure 5.

**Table 6:**
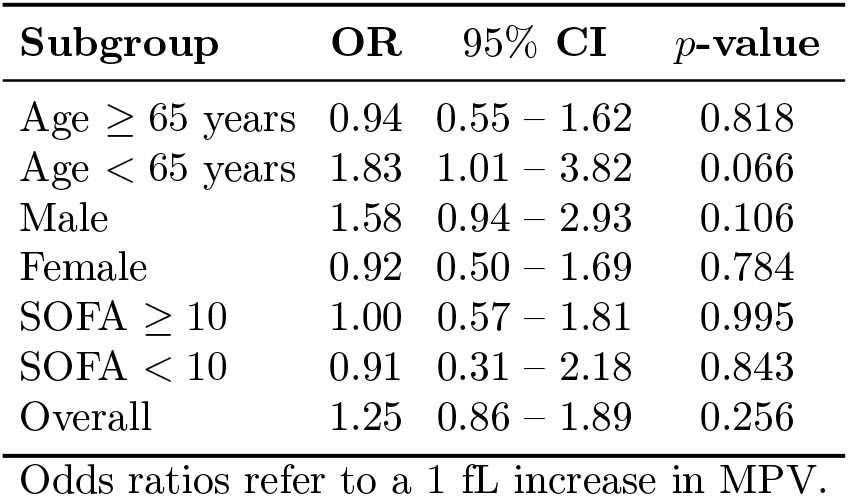
Subgroup analysis: effect of MPV on mortality.

| **Subgroup** | **OR** | **95% CI** | **p-value** |
| --- | --- | --- | --- |
| Age ≥ 65 years | 0.94 | 0.55 – 1.62 | 0.818 |
| Age < 65 years | 1.83 | 1.01 – 3.82 | 0.066 |
| Male | 1.58 | 0.94 – 2.93 | 0.106 |
| Female | 0.92 | 0.50 – 1.69 | 0.784 |
| SOFA ≥ 10 | 1.00 | 0.57 – 1.81 | 0.995 |
| SOFA < 10 | 0.91 | 0.31 – 2.18 | 0.843 |
| Overall | 1.25 | 0.86 – 1.89 | 0.256 |

Odds ratios refer to a 1 fL increase in MPV.

**Figure 5:**
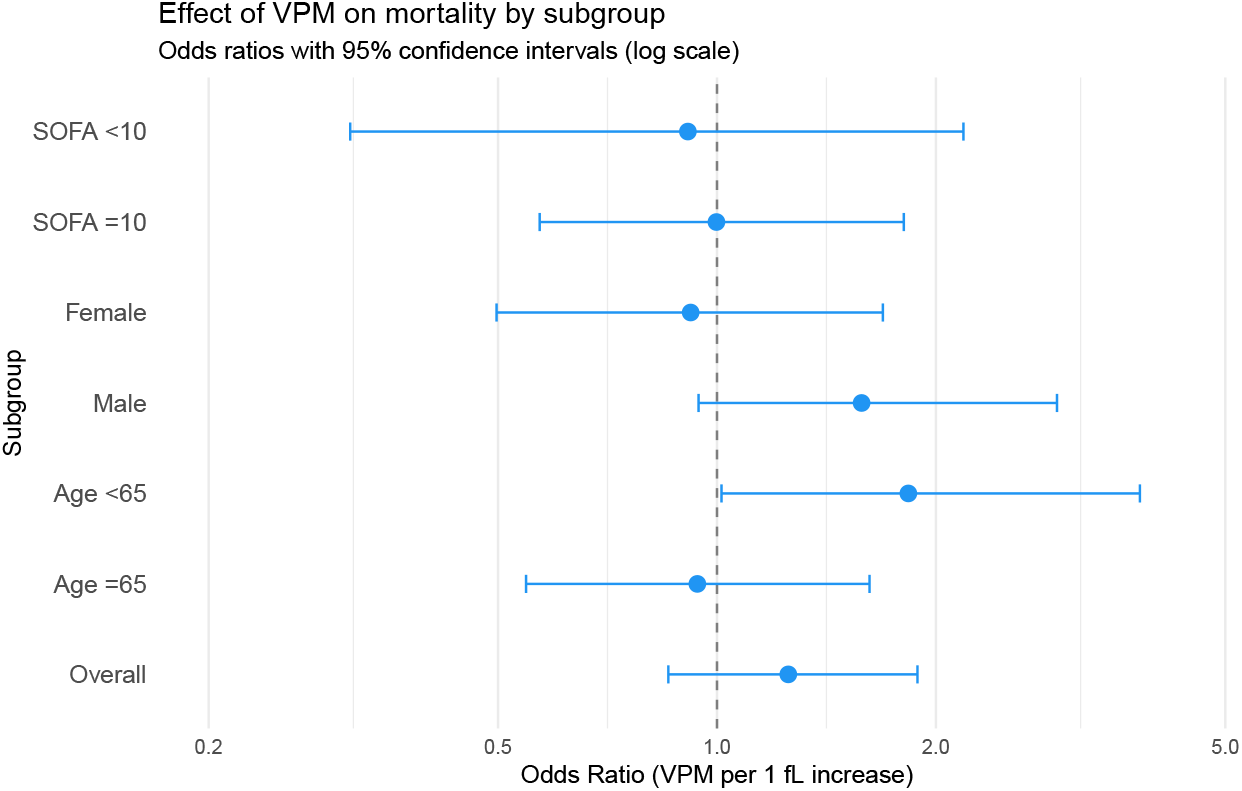
Forest plot of the effect of MPV on mortality across patient subgroups. Odds ratios (OR) with 95% confidence intervals are shown. The overall OR (1.25) and all subgroup estimates are not significant (all *p* > 0.05). A non-significant trend was observed in patients younger than 65 years (OR 1.83, 95% CI 1.01–3.82, *p* = 0.066).

### 4.1.5. Infection focus

Mortality rates did not differ significantly among infection foci (pulmonary, urinary, abdominal, soft tissue, etc.; *p* > 0.05, Chi-square test), although the study was underpowered for this comparison (data not shown). Mortality by infection focus is shown in Figure 6.

**Figure 6:**
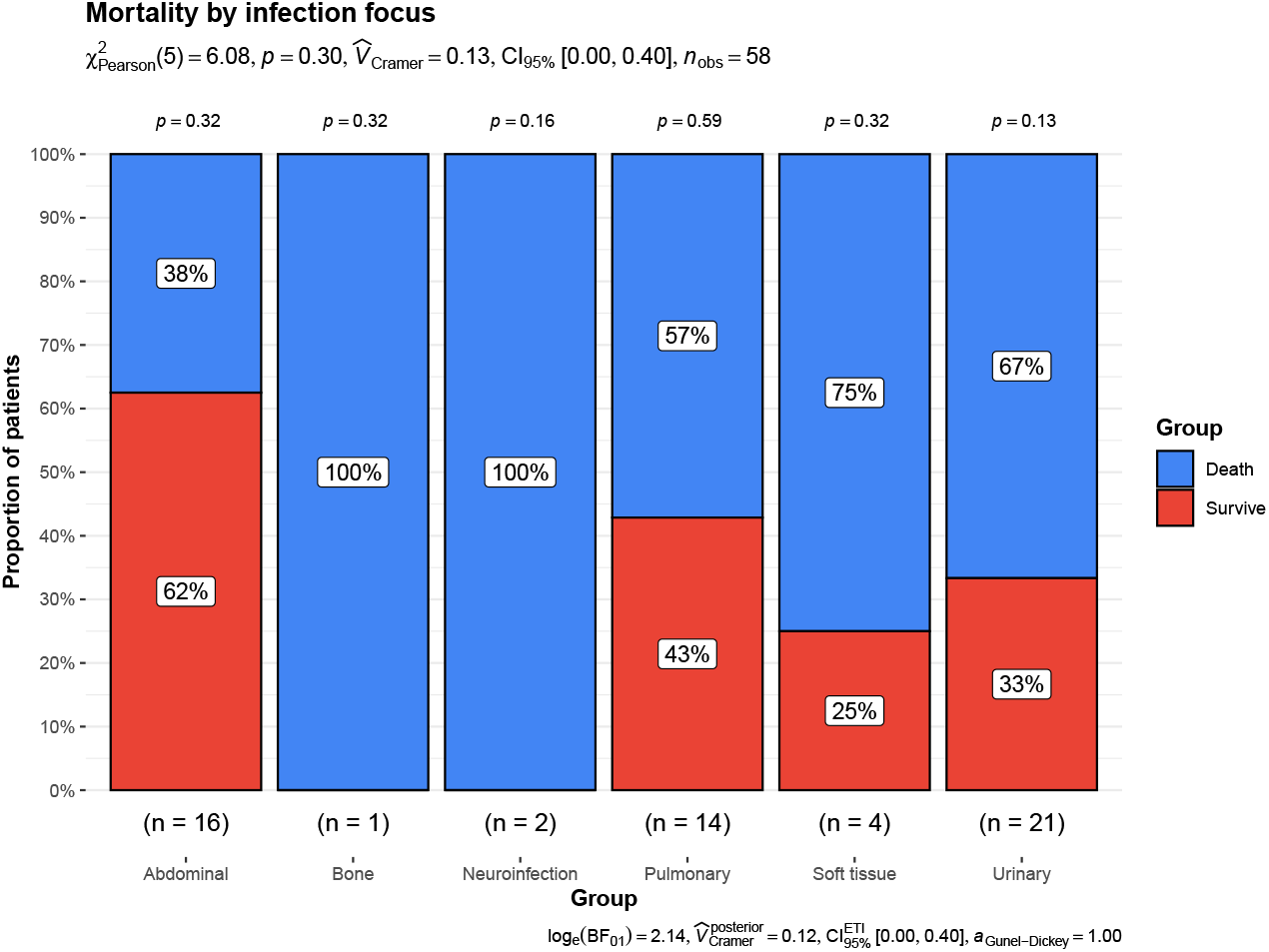
Mortality rates according to infection focus. No significant differences were observed (chi-square test, *p* > 0.05), but the study was underpowered for this comparison. Frequencies are shown as percentages (absolute numbers above bars).

## 6. Discussion

In this retrospective cohort study of 58 patients with septic shock, we found that mean platelet volume (MPV) measured at ICU admission was not associated with hospital mortality. MPV showed no significant difference between survivors and non-survivors, no independent predictive value after adjusting for disease severity, and poor discriminative ability (AUC 0.598). In contrast, conventional severity scores (SOFA, APACHE II, SAPS II) and procalcitonin performed well, with SOFA remaining the strongest independent predictor.

Our results contrast with several earlier studies that reported a positive association between elevated MPV and mortality in sepsis [9, 11]. For instance, Vélez-Páez et al. found that a MPV > 10 fL was associated with higher mortality in 64 septic patients [9]. Similarly, Zampieri et al. observed that an increase in MPV within the first 24 hours of admission predicted mortality in critically ill patients, although the cohort included mixed diagnoses [10]. However, these studies often included heterogeneous populations (sepsis without shock) and did not uniformly adjust for SOFA or other severity scores. When we restricted our analysis to septic shock and adjusted for SOFA, the effect of MPV disappeared.

The weak correlation between MPV and procalcitonin (*ρ* = 0.394, *p* = 0.002) suggests that higher MPV may reflect a more intense inflammatory response, but this relationship is modest and does not translate into improved mortality prediction. Indeed, procalcitonin itself had an AUC of 0.836, far superior to MPV.

Why does MPV fail to predict mortality in septic shock? One possible explanation is that in established shock, the overwhelming inflammatory and coagulopathic state overwhelms any subtle pro-thrombotic signal from platelet size. Moreover, MPV is influenced by many factors (e.g., thrombocytopenia, bone marrow response, timing of measurement) and may not capture the dynamic changes occurring during the first days of ICU stay [6]. Previous work has shown that changes in MPV over time, rather than a single admission value, might be more informative [10].

### 6.1. Strengths and limitations

Our study has several strengths. We used a homogeneous cohort of septic shock patients defined by current Sepsis-3 criteria [1], ensuring clinical relevance. We performed rigorous statistical analyses, including Firth’s logistic regression to address perfect separation, multivariate adjustment for key confounders, ROC comparisons, and subgroup analyses. The single-centre design allowed consistent data collection.

Limitations include the retrospective design, which carries inherent selection and information bias. The sample size is modest (58 patients), limiting power to detect small effects or interactions; however, the effect estimates were close to 1 with wide confidence intervals, suggesting no clinically meaningful association. We did not measure serial MPV values, so we cannot comment on dynamic changes. Finally, the lack of external validation precludes generalisation; our findings should be confirmed in larger, prospective, multicentre studies.

### 6.2. Clinical implications

Our results indicate that MPV at ICU admission should not be used as a prognostic marker in septic shock. Clinicians should continue to rely on validated severity scores (SOFA) and established biomarkers (procalcitonin) for risk stratification. Future research should explore whether dynamic MPV trends or combination with other platelet indices (e.g., platelet count, MPV/platelet ratio) might offer incremental value.

## 7. Conclusion

In this single-centre retrospective cohort of adult patients with septic shock, mean platelet volume measured at ICU admission was not associated with in-hospital mortality and had poor discriminatory ability (AUC 0.598). The best predictor of death was the SOFA score. MPV showed only a weak correlation with procalcitonin and no independent prognostic value after adjustment for disease severity. Subgroup analyses did not reveal any significant effect of MPV on mortality. Therefore, MPV cannot be recommended as a prognostic biomarker in septic shock. Clinicians should continue to rely on validated severity scores and established biomarkers such as procalcitonin. Larger, prospective, multicentre studies are needed to confirm these findings and to explore whether serial MPV measurements or other platelet indices might offer clinical utility.

## 8. Authors’ Contributions

FTV: conceptualization, data curation, formal analysis, funding acquisition, investigation, methodology, project administration, resources, supervision, validation, writing-reviewing and editing; PALD: data curation, formal analysis, methodology, project administration, software, supervision, validation, visualization, writing-reviewing and editing.

All authors have read and approved the final manuscript.

## 9. Acknowledgments

The authors acknowledge the *Instituto de Seguridad y Servicios Sociales de los Trabajadores del Estado Hospital Regional “General Ignacio Zaragoza”*, the *Universidad La Salle*, and the *Universidad Nacional Autónoma de México*.

## 12. Data Availability

Deidentified individual participant data will be made available following article publication, on GitHub <https://github.com/phabel-LD>, along with all analysis code.

